# Navigation behavior during visual wayfinding in people with ultra-low vision using virtual reality

**DOI:** 10.64898/2026.08.11.26360090

**Authors:** Dinesh Venugopal, Batuhan Erkat, Roksana Sadeghi, Chau Tran, Will Gee, Brittnee Livingston, Gislin Dagnelie, Arathy Kartha

## Abstract

Visual wayfinding is essential for safe navigation but remains poorly characterized in people with ultra-low vision (ULV). Because assessing complex environments in the real world carries safety risks, this study utilized a calibrated virtual reality (VR) platform to safely quantify navigation. Participants with ULV, normal vision (NV), and simulated ULV (sULV) completed tasks across three environments (street crossing, cafeteria, and metro station) of increasing complexity to determine which metrics best capture task difficulty. Navigation metrics included motion onset latency, walking speed, path efficiency, and turn deviation derived from head position data. Participants with ULV showed longer onset latency, slower walking speed, reduced path efficiency, and greater turn deviation compared with NV, while sULV showed intermediate performance. These metrics successfully reflected increasing task difficulty across environments, with the metro station posing the greatest challenge. Path efficiency consistently detected differences between environments across groups, whereas turn deviation provided insight into complex tasks. Findings indicate that diverse virtual environments capture distinct aspects of navigation that cannot be safely studied in the real world, and trajectory-based metrics capture navigation behavior more effectively than conventional measures. VR-based assessment offers a useful approach for evaluating functional navigation and guiding rehabilitation strategies in profound vision loss.

## INTRODUCTION

Independent wayfinding is essential for safe mobility, autonomy, and participation in everyday life. For people with ultra-low vision (ULV), including those with profound visual impairment or near-total blindness, navigating real-world environments can be difficult (Adeyemo et al., 2017; Geruschat, Bittner, & Dagnelie, 2012; Jeter et al., 2017; Stronks & Dagnelie, 2014). Wayfinding involves more than avoiding obstacles. It also requires understanding spatial layout, selecting an appropriate route, and making timely movement decisions when visual information is limited and often changing. As visual information is degraded in ULV, these demands place a greater cognitive burden on the person for safe and efficient navigation.

Individuals with ULV still use their rudimentary vision for a wide range of activities, including many related to mobility and gathering information about their surroundings (Adeyemo et al., 2017). Such challenges are especially pronounced in public settings such as street crossings, indoor communal spaces, and transit locations, where clutter, moving people and objects, and variable environmental conditions can increase uncertainty and compromise navigation efficiency (Kartha, Sadeghi, Swanson, & Dagnelie, 2022; Ricci, Boldini, Beheshti, Rizzo, & Porfiri, 2023). Understanding how people with ULV navigate these complex environments is therefore critical for improving mobility assessment and for informing the design of safer and more accessible navigation conditions/environments.

Studying visual wayfinding in real-world environments is difficult, particularly for people with ULV, because such studies can raise practical and safety concerns. Everyday navigation takes place in settings that are hard to control experimentally, where obstacle layout, crowd movement, traffic flow, and other features can vary across trials. When vision is degraded, navigation also places greater demands on attention, because individuals must focus on staying safe while walking at the same time as interpreting their surroundings, a trade-off demonstrated in sighted participants navigating with simulated low vision (Rand, Creem-Regehr, & Thompson, 2015). Together, these factors make it hard to separate the effects of environmental complexity from other influences on navigational behavior and raise practical and safety concerns. As a result, real-world mobility assessments, while ecologically meaningful, often lack the experimental control and measurement precision needed for direct comparisons across environments and participant groups (Chang, Dillon, Deverell, Boon, & Keay, 2020; Virgili & Rubin, 2010). In addition, wayfinding depends on spatial updating, route memory, and the integration of self-motion cues over time, indicating that navigation difficulty cannot be understood solely in terms of obstacle detection (Loomis et al., 1993).

Virtual reality (VR) offers a practical way to study visual wayfinding under controlled yet realistic conditions (Aleman et al., 2021; Authie et al., 2024; Bennett et al., 2023; Kartha, Sadeghi, Swanson, & Dagnelie, 2022; Ricci, Boldini, Beheshti, Rizzo, & Porfiri, 2023; Ricci et al., 2023). By simulating everyday environments within a controlled setting, VR allows key features such as mobility course layout with obstacles, pedestrian movement, and traffic flow to be kept consistent across participants and trials. At the same time, VR enables the safe study of demanding navigation situations, including street crossing and transit use, that would be unsafe or impractical to examine systematically in the real world for individuals with profound vision loss (Bowman & Liu, 2017). Another advantage of VR is the ability to record continuous movement data, which allows navigation behavior to be quantified using measures such as movement initiation, walking speed, path efficiency, and turning behavior (Bennett et al., 2023; Ricci et al., 2023; Ricci, Liguori, Palermo, Rizzo, & Porfiri, 2024). Collectively, these features make VR well suited for comparing how navigation behavior changes across environments with different visual and cognitive demands.

Although VR has shown promise for assessing functional vision and orientation and mobility performance in people with low vision, its use for systematically characterizing visual wayfinding in ULV remains limited. Prior VR-based studies have primarily focused on feasibility, validation, or functional task performance in isolated settings, rather than systematically comparing navigation behavior across multiple ecologically distinct environments using a common set of outcome measures (Aleman et al., 2021; Authie et al., 2024; Bennett et al., 2023). This question has become increasingly important as interest has grown in developing laboratory-based mobility courses to assess functional outcome measures for emerging retinal therapies, many of which target patients with ULV (Chung et al., 2018; Finger et al., 2019; Geruschat, Bittner, & Dagnelie, 2012; Kumaran et al., 2020). The broader extended reality literature (encompassing VR, augmented reality, and other immersive mixed reality environments) also shows growing interest in applying these technologies to vision research, while noting a wide variation in tasks, outcome measures, and study design (Htike, Margrain, Lai, & Eslambolchilar, 2020; Kasowski, Johnson, Neydavood, Akkaraju, & Beyeler, 2023). Conventional mobility metrics such as completion time, obstacle collision, or course-based walking speed and percentage of preferred walking speed, may not fully capture how individuals navigate through an environment, particularly when the path deviates from the intended course because of obstacle avoidance, hesitation, or reorientation (Chang, Dillon, Deverell, Boon, & Keay, 2020; Ruiz, Garces, Soo, & Fernandez, 2025; Virgili & Rubin, 2010). As a result, it remains unclear both which environments pose the greatest challenge for individuals with ULV, and which trajectory-based measures best reflect meaningful differences in visually guided wayfinding.

The present study used a previously developed and calibrated VR wayfinding instrument, Wilmer VRW (Kartha, Sadeghi, Swanson, & Dagnelie, 2022), to examine navigation behavior in individuals with ULV, simulated ULV (sULV), and normal vision (NV) across three ecologically relevant environments: street crossing, cafeteria, and metro station. Navigation behavior was quantified using four complementary outcomes that captured different aspects of visually guided wayfinding: motion onset latency, walking speed, path efficiency, and turn deviation. We hypothesized that navigation difficulty would increase with environmental complexity, particularly in participants with ULV and sULV, relative to participants with NV, and that trajectory-based measures such as path efficiency and turn deviation would be especially informative for detecting these differences.

## METHODS

### Participants

Participants included adults with NV and ULV who were recruited from Johns Hopkins Department of Ophthalmology. The ULV group was defined as individuals with visual acuity of ≤ 20/1600 (1.9 logMAR) (Geruschat, Bittner, & Dagnelie, 2012). For the NV group, participants with no ocular disease, no history of ocular surgery and visual acuity of 20/25 (0.1 logMAR) or better in each eye. Individuals who self-reported any cognitive impairment, hearing impairment, or mobility impairment were not included in the study.

The study was approved by the Johns Hopkins Medicine Institutional Review Board. All participants were treated in accordance with the tenets of the Declaration of Helsinki and written informed consent was obtained prior to participation.

Participants with ULV and NV completed the VR visual wayfinding tasks with their habitual correction. Participants with NV also performed the VR visual wayfinding task under simulated ULV conditions. The ULV was simulated using a combination of multiple Bangerter filters that decreased visual acuity to approximately 20/2000 (2.0 logMAR).

### Visual wayfinding tasks

All visual wayfinding tasks were presented using an HTC VIVE Pro Eye headset (1440 × 1600 pixels per eye resolution, 110° diagonal field of view, 90 Hz frame rate). All participants wore the VR headset and completed ten wayfinding tasks across three VR environments in the order as specified: street crossing, cafeteria, and metro station. Each environment presented up to four difficulty levels with increasing environmental complexity. Participants were provided with a practice trial right before starting each level in the three VR environments. Head position was continuously tracked during all tasks.

#### Street Crossing

In the street crossing environment, participants were asked to complete street crossing tasks under four different difficulty levels that were presented sequentially with increasing complexity. The first level was an empty one-way street (level A), second was a one-way street with parked cars and moving avatars (acted as pedestrians) (level B), third involved two-way street with moving cars and traffic gap judgements (level C), and fourth involved everything in the third level plus moving pedestrians (level D). Figure 1 illustrates the overhead views of street crossing environments for each difficulty level. Across all four difficulty levels, participants were instructed to step down a curb, and to reach the opposite curb approximately 4.6 m away. They were instructed to follow the crosswalk and walk at a comfortable walking speed while avoiding cars and pedestrians. In the scenarios involving pedestrians, two pedestrians approached from the opposite direction while two walked alongside the participant, without obstructing the participant’s walking path.

**Figure 1.**
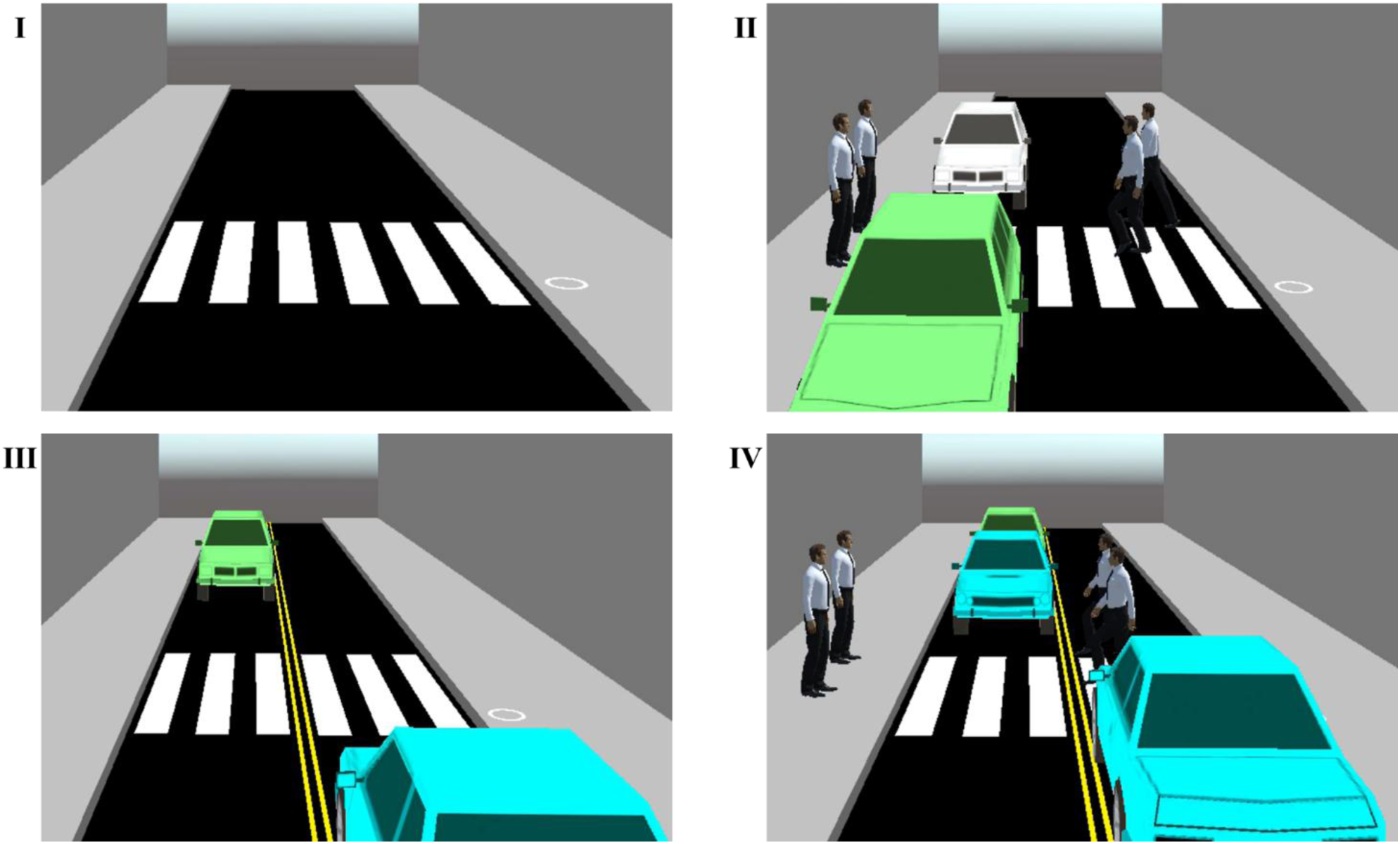
Overhead views of the street crossing environment at all four difficulty levels (panels I to IV). Panel I shows an empty one-way street (level A); panel II shows a one-way street with parked cars and moving pedestrians (level B); panel III shows a two-way street with moving cars (level C); and panel IV shows a two-way street with moving cars and moving pedestrians (level D). The white circle on the right side of the curb visible in panels I to III represents the start location of the participant in the virtual environment. In panels III and IV, the two-way streets are marked by yellow lane markings. In panels II and IV, two pedestrians are positioned halfway into the street, indicating that they walk alongside the participant, while two pedestrians on the opposite curb waiting for the participant to start walking.

#### Cafeteria

In the cafeteria environment, participants had to complete the task of finding a cashier under three difficulty levels presented sequentially with increasing complexity. Participants were asked to locate and walk to the cashier located at the end of the room, approximately 4.3 m away, for each level. The first level was tables organized in an ordered manner (level A), second was disorganized tables (level B), and third a party scene with cluttered surroundings including disorganized tables, seated avatars (acted as customers), orange balloons hanging from a few tables, posters on the walls, and a moving avatar (acted as a waiter) (level C). Figure 2 illustrates the participant’s first-person view of cafeteria environment for each difficulty level. The cashier location was randomized to the left, center, or right across levels and participants. In the two scenarios with disorganized tables, table location and orientation were randomized across participants. Across all levels, participants started walking at a comfortable speed from a predefined starting location (door entrance) and walked toward the cashier, while avoiding tables and the waiter. The waiter walked from left to right or right to left (randomized across participants) after the participant started walking, without obstructing the participant’s walking path.

**Figure 2.**
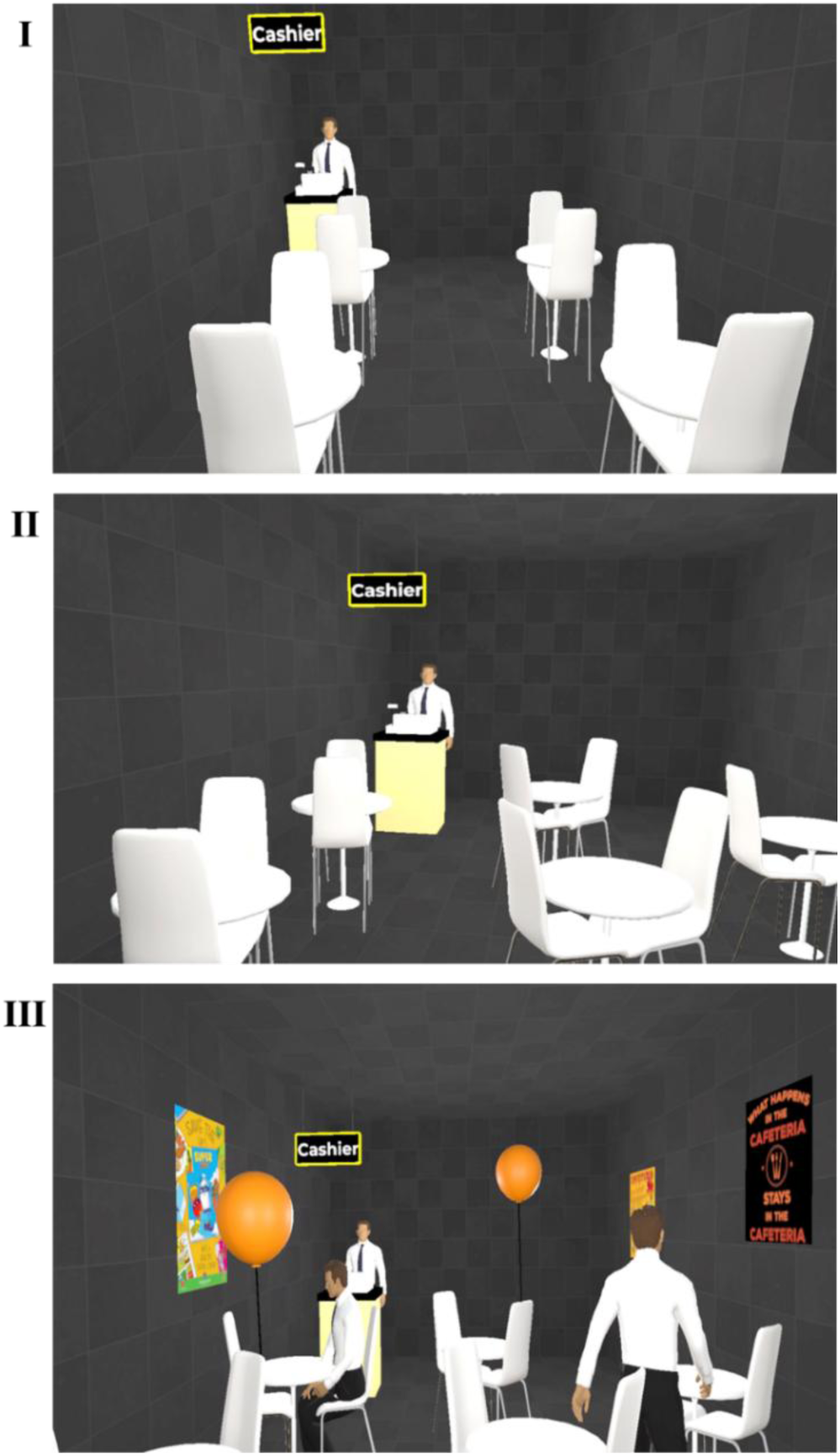
Participant’s first-person views of the cafeteria environment at all three difficulty levels (panels I to III). Panel I shows a cafeteria scene with tables organized in an ordered manner (level A); panel II shows disorganized tables (level B); and panel III shows cluttered surroundings including disorganized tables, seated customers, orange balloons hanging from a few tables, posters on the walls, and a moving waiter (level C). In these examples, the cashier is located on the left side of the cafeteria. In panel III, the waiter is positioned on the right side of the cafeteria, indicating that he walks from left to right.

#### Metro Station

In the metro station environment, participants had to complete the task of boarding a metro under three difficulty levels presented sequentially with increasing complexity. Across all difficulty levels, at first, the participant was standing at a curb in the metro platform (the final step of a staircase), then stepped down, and started walking along the platform at a comfortable walking speed. In the first level, participants were asked to locate the train entrance door by positioning themselves behind the yellow line on the platform (level A). There was no train arriving at this level. In the second level, participants had to complete steps from level one and then enter the train with avatars around (acted as passengers) (level B). In the third level, participants had to complete all the steps in the second level and afterwards identify an empty seat inside the train (level C). Figure 3 illustrates the participant’s first-person view of metro station environment for three difficulty levels. Participants were instructed to avoid bumping into the passengers while performing the task. The location of the train entrance door was randomized across levels and participants (1.2 m, 2.5 m, or 4.0 m to the right of the starting position). While entering the train, one passenger walked alongside the participant and two passengers exited the train, without obstructing the participant’s walking path. Participants were asked to wait for the exiting passengers to leave the train before boarding the train. The train always arrived from right to left as the participant reached the entrance door in the second and third levels (levels B and C).

**Figure 3.**
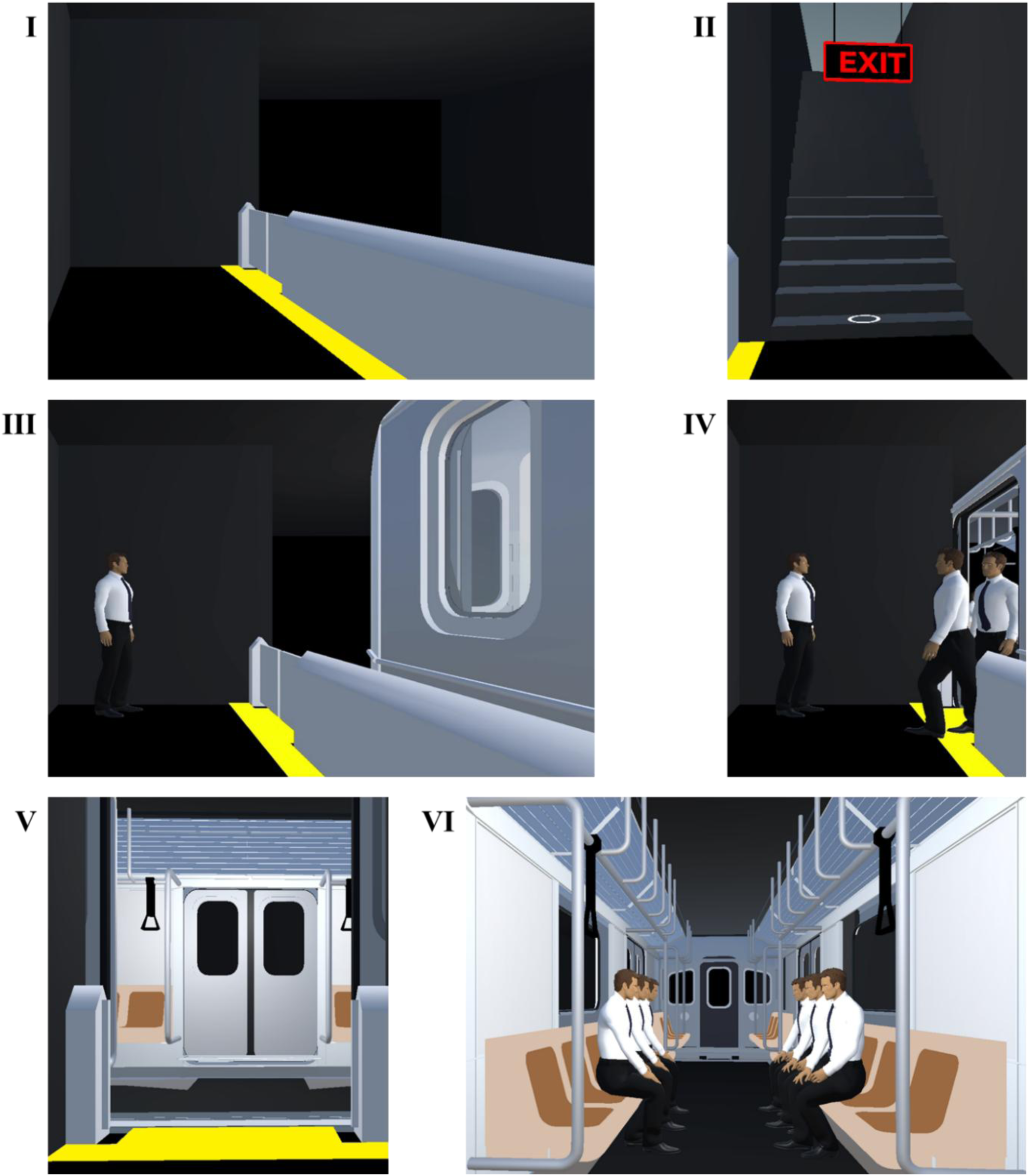
Participant’s first-person views of the metro station environment at all three difficulty levels (panels I to VI). Panel I shows the train entrance door (level A). Panel II shows the start location of the participant in the virtual environment, indicated by a white circle on the final step of a stairway (levels A, B, and C). Panel III shows the train entrance door, a waiting passenger, and the metro train arriving from right to left (levels B and C). Panel IV shows the train entrance door, a waiting passenger, the metro train, and two passengers exiting the train (levels B and C). Panel V shows the train entrance door (levels B and C). Panel VI shows the empty seats inside the train on the right and left sides, with passengers seated around (level C).

**Figure 4.**
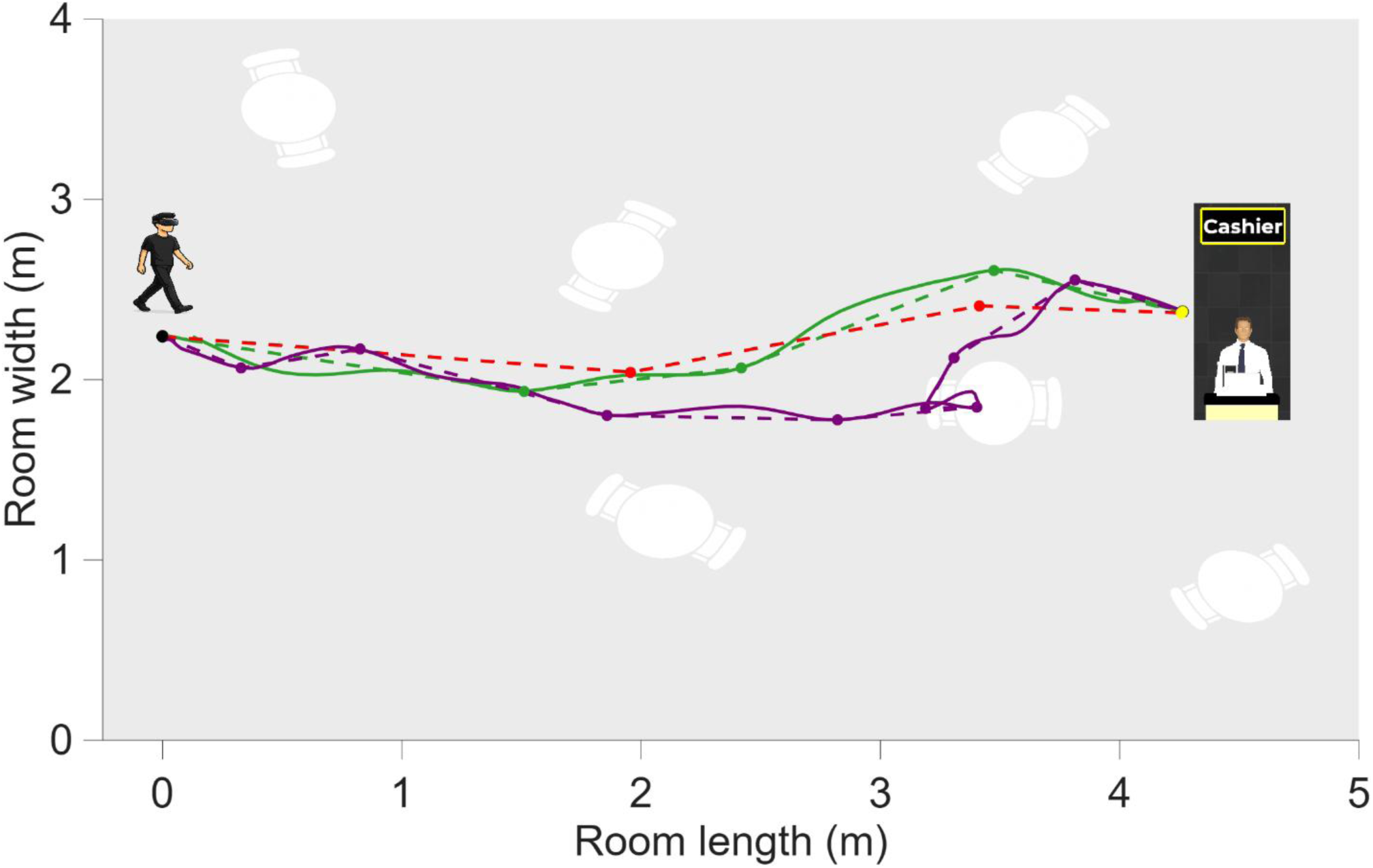
Bird’s-eye view of the walking paths in the cafeteria environment with disorganized tables (level B). The black dot represents the start location, and the yellow dot represents the cashier location. The path walked by a participant with ultra-low vision is shown as a purple solid line, with the corresponding simplified Ramer-Douglas-Peucker segmented path shown as a purple dashed line and purple dots indicating turns. The path walked by a participant with normal vision is shown as a green solid line, with the corresponding simplified Ramer-Douglas-Peucker segmented path shown as a green dashed line and green dots indicating turns. Both participant paths are compared with the optimal path (red dashed line) and expected turns (red dots). The solid purple line indicates that a participant with ultra-low vision collided with the table obstacle in the virtual environment.

Performance was assessed using only visual cues, there were no auditory or tactile feedback (except a wooden plank used for curbs for street crossing and a staircase step for metro station). The location of the wooden plank was matched with their location in VR environments.

### Head tracking

Head position data were collected as vectors for all three environments and exported to MATLAB for analysis. Data included time-stamped three-dimensional positional data along the X (forward), Y (vertical), and Z (lateral) axes and processed in MATLAB (R2025b; MathWorks, Natick, MA, USA) for each VR wayfinding task. Note that head orientation data were not included in this analysis; they are being extracted and will be analyzed for possible future publication.

### Data analysis

The first step was to process the raw head position vectors. Data were arranged sequentially based on their recorded time-stamps. Tasks with missing data were excluded. The analyses were limited to the data captured between the starting position and task completion. Any data after the task was completed were excluded. After processing, head position data were used to derive path efficiency, turn deviation and motion onset latency.

#### Path efficiency

Given that the observed walking paths varied between environments, complexity levels and participants, it was necessary to define the optimal path. This optimal path was then compared with the observed path of the participants. Optimal path was defined in MATLAB by manually selecting waypoints representing the shortest feasible route between the starting position and task completion while avoiding obstacles. From these interactive markings, the optimal walking path, optimal path length, and the number of intentional turns required to complete the task were derived. Intentional turns were defined as discrete changes in walking direction between successive segments of the optimal path. These turns indicated intentional reorientation during navigation rather than minor positional fluctuations and provided a descriptive measure of path structure and directional planning during wayfinding. To assess the reproducibility of optimal path marking, two independent observers (DV and BE) performed these markings on a subset of 20 instances. Agreement between observers was evaluated using Bland-Altman analysis. The mean ± standard deviation (95% limits of agreement) of the difference in optimal path distance and optimal turn counts between two observers were 0.03 ± 0.12 m (0.26 to −0.21) and 0.2 ± 0.5 turns (1.1 to −0.8) respectively, demonstrating good agreement between observers. Path efficiency was then calculated as the ratio of optimal path length to observed distance walked, reported as percentage (%), with higher values indicating more efficient navigation.

#### Turn deviation

Turn deviation was derived from simplified walking path and the optimal path. The simplified walking path was obtained from observed walking path using the simplified Ramer-Douglas-Peucker (RDP) algorithm (ε = 0.1 m) to reduce noise while preserving major directional changes (Douglas & Peucker, 1973; Ramer, 1972). The number of retained RDP points, excluding the starting position and task completion, were considered as the number of turns in the observed path (Bertaud et al., 2021). Turn deviation was calculated as the difference between the number of turns in the observed (RDP-derived) path and the number of intentional turns in the optimal path.

#### Motion onset latency

Motion onset latency (in seconds, s) was based on two-stage criteria. The primary criterion was “a curb step-down event”, operationally defined as the first occurrence of a vertical head position drop of at least 0.1 m occurring within a forward displacement window of 0.5 m from the starting position. If this criterion was met, motion onset latency was calculated as the time elapsed between start and the detected curb step-down event. If primary criterion was not met (e.g., in cafeteria wayfinding tasks, where a curb was not present), a secondary criterion “walking initiation event” was applied to identify motion onset latency. Walking initiation was defined as the first instance in which forward displacement exceeded 0.3 m relative to the starting position. Here, motion onset latency was calculated as the time elapsed from start to the walking initiation event. For each task, motion onset latency, and the criterion with which latency was derived were recorded. In the current study, 95% of street crossing and 92% of metro station wayfinding tasks were obtained from the primary criterion, with the remaining tasks derived from the secondary criterion. Smaller motion onset latency values reflect faster initiation of walking.

Active task duration (in seconds, s) was defined as the elapsed time from motion onset to task completion and was calculated by subtracting motion onset latency from the total task duration. Walking speed (in meters per second, m/s) for each task was calculated as the observed walking distance divided by the active task duration, using head position and time-stamp data. Therefore, walking speed reflected movement speed after task initiation and did not include the initial decision or waiting period. Task completion time and collision events were reported elsewhere (Kartha, Sadeghi, Swanson, & Dagnelie, 2022).

Statistical analyses were conducted using IBM SPSS Statistics for Windows (version 27.0; IBM Corp., Armonk, NY, USA), with a significance level set at p < 0.05. Generalized linear mixed models with Bonferroni-adjusted post-hoc pairwise comparisons were used to compare outcome measures between the NV, sULV, and ULV groups, with separate models fitted for each tested environment (street crossing, cafeteria, and metro station). Fixed effects included participant group (NV, sULV, ULV), level within each environment, and their interaction. Participant was included as a random effect to account for repeated measures across tasks. For post-hoc comparisons, the Bonferroni-adjusted significance threshold was α = 0.017 for three pairwise group comparisons and for three pairwise level comparisons, and α = 0.008 for six pairwise comparisons among four levels. To determine which environment elicited more difficulty, we collapsed all difficulty levels (except level A of the street crossing environment, which contained no obstacles and required walking straight ahead) and compared wayfinding performance across street crossing, cafeteria and metro station within each participant group. A three-way repeated measures analysis of variance, with Bonferroni-adjusted post-hoc pairwise comparisons, was used to compare the tasks across environments. To explore individual variability, using these collapsed measures, Pearson correlation coefficients were calculated within the ULV group to examine the relationship between visual acuity and each of the trajectory-based navigation metrics separately for each environment.

## RESULTS

### Participant characteristics

Fifteen participants with ULV and ten participants with normal vision (NV) participated in the study. Details related to diagnosis of ULV participants are summarized in Table 1. The mean age of participants with NV was 35.1 years (SD: 10.5 years, Female: 50%). Head position data were unavailable for all ten tasks completed by one participant with NV. Additionally, in a small number of instances, completion times were missing, which prevented computation of motion onset latency (in metro station environment: 3%, 3/99), walking speed (street crossing environment: 3%, 4/136; metro station environment: 5%, 5/99), path efficiency (street crossing environment: 3%, 4/136; metro station environment: 5%, 5/99), and turn deviation (street crossing environment: 3%, 4/136; metro station environment: 5%, 5/99). Figure 5 illustrates bird’s-eye view of walking paths from example participants with ULV and NV in the cafeteria environment, with corresponding simplified RDP-segmented paths and turns, compared with the optimal path at level B.

**Figure 5.**
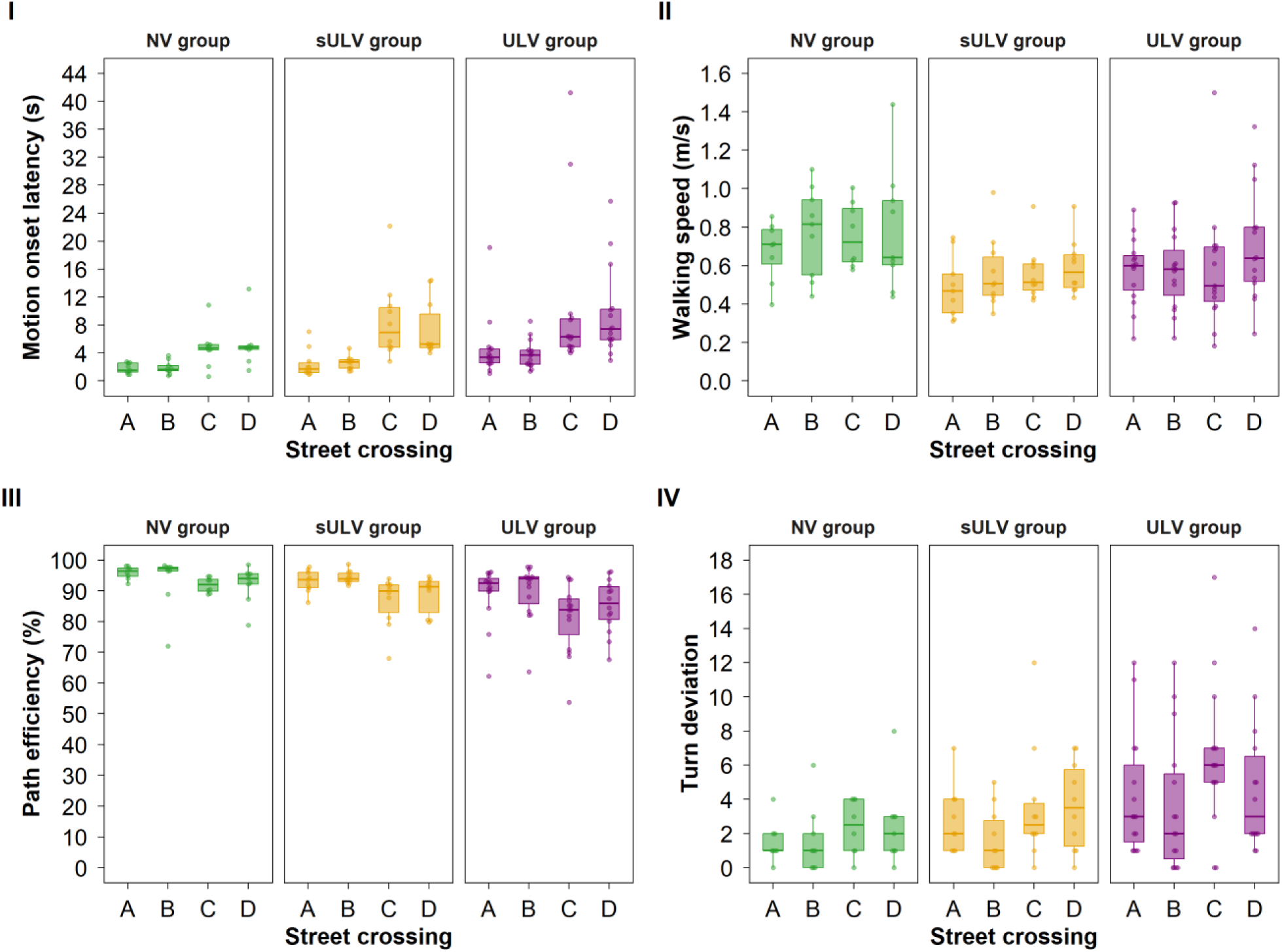
Box plots showing motion onset latency (I), walking speed (II), path efficiency (III) and turn deviations (IV) for the four street crossing tasks (A. empty street, B. street with parked cars and moving pedestrians, C. moving cars, and D. moving cars with moving pedestrians) and the three groups (normal vision [NV], simulated ultra-low vision [sULV], and ultra-low vision [ULV]). Across all panels, box plots and overlaid data points use the same color, with NV shown in green, sULV in amber, and ULV in purple. The box indicates the interquartile range (25^th^-75^th^ percentile) and median; the whiskers indicate the minimum and maximum values excluding outliers.

**Table 1.**
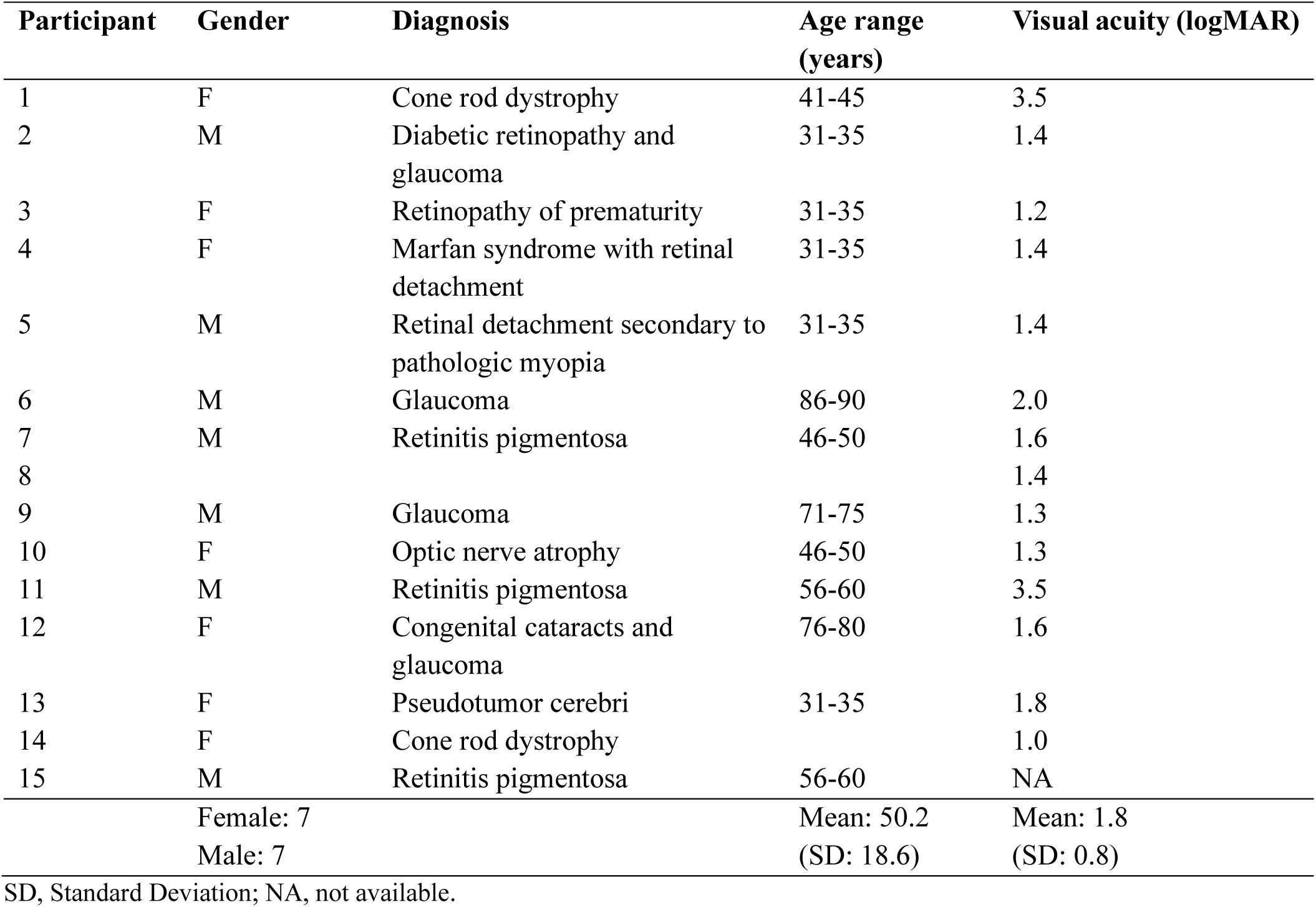
The descriptive information about each participant with ultra-low vision.

#### Street Crossing

Motion onset latency, walking speed, path efficiency, and turn deviation across the four street crossing tasks are shown in Figure 5.

For motion onset latency in the street crossing tasks, there was a significant main effect of participant group (F(2,124) = 4.71, p = 0.011), with the ULV group initiating movement 3.6 s later than the NV group (7.0 vs. 3.4 s, approximately 2.1 times as long as the NV group, p = 0.003). A significant main effect of level was also noted (F(3,124) = 10.33, p < 0.001). Post-hoc analysis revealed that the moving cars (level C) and moving cars with moving pedestrians (level D) delayed walking initiation by 4.9 s (7.8 vs. 2.9 s, approximately 2.7 times as long, p < 0.001) and 4.4 s (7.4 vs 2.9 s, approximately 2.5 times as long, p < 0.001), respectively, compared with the empty street level (A). However, there was no significant interaction between participant group and level (F(6,124) = 0.36, p = 0.904, Figure 5.I).

Walking speed did not differ significantly across participant groups (F(2,120) = 2.41, p = 0.094), and there were no significant group by level interaction (F(6,120) = 0.52, p = 0.792, Figure 5.II). However, a significant main effect of level was found (F(3,120) = 3.62, p = 0.015), with moving cars with moving pedestrians (level D) showing higher walking speed by 0.12 m/s compared with the empty street level (A) (0.69 vs. 0.57 m/s, p = 0.001), averaged across all participant groups. This finding should be interpreted cautiously because walking speed was calculated after removing motion onset latency and therefore reflects movement speed after crossing initiation rather than overall task speed.

For path efficiency, a significant main effect was found for participant group (F(2,120) = 3.70, p = 0.028), with the ULV group performing 7.2% less efficiently than the NV group (p = 0.011), and no differences were observed between the NV and sULV groups (p = 0.403) or between the sULV and ULV groups (p = 0.084). A significant level effect was also noted (F(3,120) = 11.58, p < 0.001), with reduced path efficiency for moving cars (level C) and moving cars with moving pedestrians (level D) by 6.1% (p < 0.001) and 4.3% (p = 0.001), respectively, compared to empty street level (A). Nevertheless, there was no significant group by level interaction effect (F(6,120) = 0.89, p = 0.506, Figure 5.III).

Turn deviation showed a significant main effect of participant group (F(2,120) = 3.09, p = 0.049), with the ULV group making 3 more turns than the NV group (p = 0.02), and a significant level effect (F(3,120) = 6.86, p < 0.001), with moving cars (level C) requiring 1 more turn compared with the empty street level (A) (p = 0.012). However, no significant interaction between group and level was observed (F(6,120) = 0.76, p = 0.606, Figure 5.IV).

Generalized linear mixed models results for all outcome measures of street crossing tasks, including F values, degrees of freedom, and p value, are provided in Table 2. Bonferroni-adjusted post-hoc pairwise comparisons between the three participant groups for all street crossing outcome measures are summarized in Table 3.

**Table 2.** Generalized linear mixed models for all the outcome measures of street crossing tasks.

| Outcome measures | Effects | F value | df1 | df2 | p value |
| --- | --- | --- | --- | --- | --- |
| Motion onset latency | Group | 4.71 | 2 | 124 | **0.011** |
|  | Level | 10.33 | 3 | 124 | **< 0.001** |
|  | Group X Level | 0.36 | 6 | 124 | 0.904 |
| Walking speed | Group | 2.41 | 2 | 120 | 0.094 |
|  | Level | 3.62 | 3 | 120 | **0.015** |
|  | Group X Level | 0.52 | 6 | 120 | 0.792 |
| Path efficiency | Group | 3.70 | 2 | 120 | **0.028** |
|  | Level | 11.58 | 3 | 120 | **< 0.001** |
|  | Group X Level | 0.89 | 6 | 120 | 0.506 |
| Turn deviation | Group | 3.09 | 2 | 120 | **0.049** |
|  | Level | 6.86 | 3 | 120 | **< 0.001** |
|  | Group X Level | 0.76 | 6 | 120 | 0.606 |

Significant p values are indicated in bold.

**Table 3.** Bonferroni-adjusted post-hoc pairwise comparisons between the three groups for all outcome measures of street crossing environment.

|  |  | Mean difference (95% confidence interval) |  |
| --- | --- | --- | --- |
| Motion onset latency (s) |  | NV group | sULV group |
| A | ULV group | 2.79 (-1.44 to 7.03) | 2.12 (-1.98 to 6.22) |
|  | sULV group | 0.68 (-3.94 to 5.29) | — |
| B | ULV group | 1.91 (-2.33 to 6.14) | 1.23 (-2.87 to 5.33) |
|  | sULV group | 0.68 (-3.94 to 5.29) | — |
| C | ULV group | **5.40 (1.17 to 9.63)** | 1.53 (-2.57 to 5.63) |
|  | sULV group | 3.87 (-0.75 to 8.48) | — |
| D | ULV group | 4.43 (0.20 to 8.67) | 2.21 (-1.89 to 6.31) |
|  | sULV group | 2.22 (-2.39 to 6.84) | — |
| Walking speed (m/s) |  |  |  |
| A | ULV group | -0.07 (-0.27 to 0.12) | 0.10 (-0.08 to 0.29) |
|  | sULV group | -0.18 (-0.39 to 0.04) | — |
| B | ULV group | -0.20 (-0.39 to -0.01) | 0.02 (-0.17 to 0.20) |
|  | sULV group | -0.22 (-0.42 to -0.01) | — |
| C | ULV group | -0.15 (-0.34 to 0.04) | 0.02 (-0.16 to 0.21) |
|  | sULV group | -0.17 (-0.38 to 0.04) | — |
| D | ULV group | -0.10 (-0.29 to 0.09) | 0.09 (-0.09 to 0.27) |
|  | sULV group | -0.19 (-0.39 to 0.02) | — |
| Path efficiency (%) |  |  |  |
| A | ULV group | -6.32 (-13.04 to 0.40) | -3.67 (-10.15 to 2.81) |
|  | sULV group | -2.65 (-10.04 to 4.75) | — |
| B | ULV group | -3.78 (-10.36 to 2.79) | -4.58 (-10.95 to 1.79) |
|  | sULV group | 0.80 (-6.37 to 7.96) | — |
| C | ULV group | **-10.12 (-16.84 to -3.39)** | -5.27 (-11.64 to 1.10) |
|  | sULV group | -4.85 (-12.15 to 2.45) | — |
| D | ULV group | **-8.57 (-15.20 to -1.95)** | -5.15 (-11.56 to 1.27) |
|  | sULV group | -3.43 (-10.59 to 3.74) | — |
| Turn deviation |  |  |  |
| A | ULV group | 2.50 (-0.23 to 5.23) | 1.16 (-1.47 to 3.79) |
|  | sULV group | 1.34 (-1.66 to 4.34) | — |
| B | ULV group | 2.04 (-0.64 to 4.73) | 2.00 (-0.60 to 4.60) |
|  | sULV group | 0.04 (-2.88 to 2.97) | — |
| C | ULV group | **3.82 (1.10 to 6.55)** | 2.80 (0.20 to 5.40) |
|  | sULV group | 1.02 (-1.94 to 3.99) | — |
| D | ULV group | 2.75 (0.05 to 5.44) | 1.59 (-1.02 to 4.20) |
|  | sULV group | 1.16 (-1.77 to 4.08) | — |

Significant mean differences (horizontal subtracted by vertical) are indicated in bold.

#### Cafeteria

Figure 6 demonstrates motion onset latency, walking speed, path efficiency, and turn deviation across the three cafeteria environment wayfinding tasks for the three participant groups.

**Figure 6.**
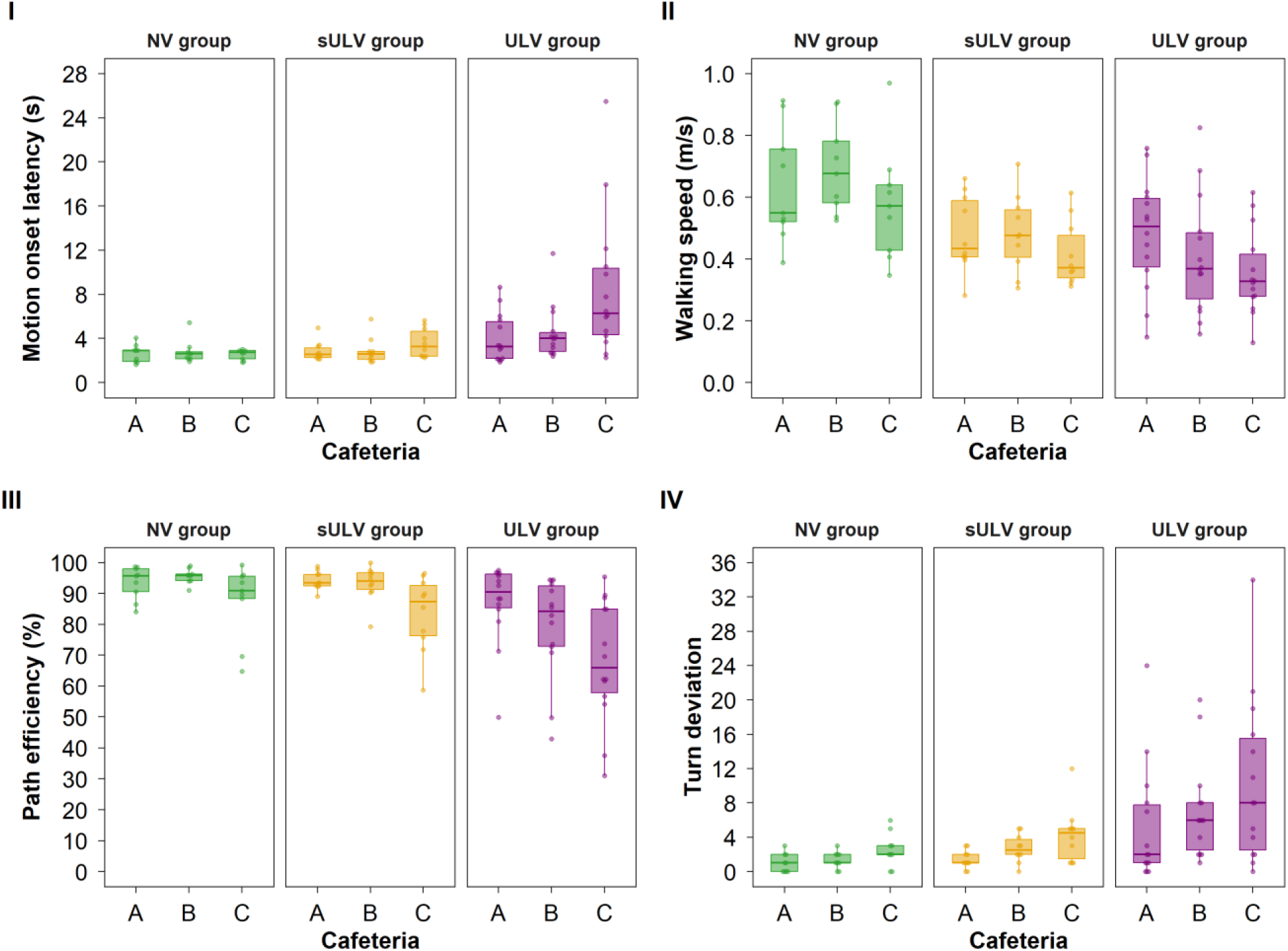
Box plots showing motion onset latency (I), walking speed (II), path efficiency (III) and turn deviations (IV) for the three cafeteria tasks (A. organized tables, B. disorganized tables, and C. cluttered surroundings) and the three groups (normal vision [NV], simulated ultra-low vision [sULV], and ultra-low vision [ULV]). Across all panels, box plots and overlaid data points use the same color, with NV shown in green, sULV in amber, and ULV in purple. The box indicates the interquartile range (25^th^-75^th^ percentile) and median; the whiskers indicate the minimum and maximum values excluding outliers.

In the cafeteria environment wayfinding tasks, onset latency showed significant main effects of participant group (F(2,90) = 6.79, p = 0.002) and level (F(2,90) = 5.43, p = 0.006), as well as a significant interaction effect (F(4,90) = 4.27, p = 0.003, Figure 6.I). Post-hoc analysis revealed that in the cluttered surroundings (level C), the ULV group initiated walking 6.0 s and 5.0 s later than the NV and sULV groups (8.6 vs. 2.6 s, approximately 3.3 times as long as the NV group, p < 0.001; and 8.6 vs. 3.6 s, approximately 2.4 times as long, p < 0.001), respectively, while no difference was found between NV and sULV groups (p = 0.438). Additionally, no significant differences were observed between the organized and disorganized tables (levels A and B) across participant groups (p > 0.017).

Walking speed showed a significant main effect of participant groups (F(2,90) = 8.20, p = 0.001), with the ULV and sULV walking 0.22 m/s and 0.18 m/s slower than the NV group (0.42 vs. 0.64 m/s, approximately 0.6 times the speed of the NV group, p < 0.001; and 0.46 vs. 0.64 m/s, approximately 0.7 times the speed, p = 0.004, respectively). There was also a significant level effect (F(2,90) = 5.99, p = 0.004), where the cluttered surroundings (level C) reduced walking speed by 0.08 m/s relative to both organized (level A) and disorganized tables tasks (level B) (0.45 vs. 0.53 m/s, approximately 0.8 times the speed of level A, p = 0.003; and 0.45 vs. 0.53 m/s, approximately 0.8 times the speed of level B, p = 0.004). No significant interaction between level and group was observed (F(4,90) = 1.02, p = 0.401, Figure 6.II).

Path efficiency showed significant main effects of participant group (F(2,90) = 5.87, p = 0.004) and level (F(2,90) = 23.94, p < 0.001), as well as significant interaction (F(4,90) = 2.57, p = 0.043, Figure 6.III). Post-hoc analysis revealed that in both the disorganized tables (level B) and cluttered surroundings (level C), participants with ULV performed 16.1% and 19.5% less efficiently than the NV (p = 0.003 and p < 0.001, respectively). They also performed 13.7% and 15.4% less efficiently than the sULV group (p = 0.009 and p = 0.003, respectively). No significant difference was found between the NV and the sULV groups across all three cafeteria tasks (p > 0.017).

For turn deviation, a significant main effect was observed for participant groups (F(2,90) = 5.24, p = 0.007), with the ULV requiring 6 more turns than the NV group (p = 0.004). There was also a significant level effect (F(2,90) = 11.48, p < 0.001), where the cluttered surroundings (level C) elicited 3 and 2 more turns than the organized (level A) (p < 0.001) and disorganized tables (level B) (p = 0.003), respectively. However, no significant interaction effect between group and level was observed (F(4,90) = 1.34, p = 0.262, Figure 6.IV).

Generalized linear mixed models results for all outcome measures of cafeteria environment wayfinding tasks, including F values, degrees of freedom, and p value, are provided in Table 4. Bonferroni-adjusted post-hoc pairwise comparisons between the three participant groups for all cafeteria environment outcome measures are summarized in Table 5.

**Table 4.** Generalized linear mixed models for all the outcome measures of cafeteria environment wayfinding tasks.

| Outcome measures | Effects | F value | df1 | df2 | p value |
| --- | --- | --- | --- | --- | --- |
| Motion onset latency | Group | 6.79 | 2 | 90 | **0.002** |
|  | Level | 5.43 | 2 | 90 | **0.006** |
|  | Group X Level | 4.27 | 4 | 90 | **0.003** |
| Walking speed | Group | 8.20 | 2 | 90 | **0.001** |
|  | Level | 5.99 | 2 | 90 | **0.004** |
|  | Group X Level | 1.02 | 4 | 90 | 0.401 |
| Path efficiency | Group | 5.87 | 2 | 90 | **0.004** |
|  | Level | 23.94 | 2 | 90 | **< 0.001** |
|  | Group X Level | 2.57 | 4 | 90 | **0.043** |
| Turn deviation | Group | 5.24 | 2 | 90 | **0.007** |
|  | Level | 11.48 | 2 | 90 | **< 0.001** |
|  | Group X Level | 1.34 | 4 | 90 | 0.262 |

Significant p values are indicated in bold.

**Table 5.** Bonferroni-adjusted post-hoc pairwise comparisons between the three groups for all outcome measures of cafeteria environment.

|  |  | Mean difference (95% confidence interval) |  |
| --- | --- | --- | --- |
| Motion onset latency (s) |  | NV group | sULV group |
| A | ULV group | 1.38 (-1.05 to 3.81) | 1.17 (-1.18 to 3.53) |
|  | sULV group | 0.21 (-2.40 to 2.83) | — |
| B | ULV group | 1.72 (-0.72 to 4.15) | 1.65 (-0.71 to 4.00) |
|  | sULV group | 0.07 (-2.55 to 2.68) | — |
| C | ULV group | **6.00 (3.57 to 8.43)** | **4.98 (2.62 to 7.33)** |
|  | sULV group | 1.03 (-1.59 to 3.64) | — |
| Walking speed (m/s) |  |  |  |
| A | ULV group | -0.16 (-0.29 to -0.02) | 0.00 (-0.13 to 0.13) |
|  | sULV group | -0.16 (-0.30 to -0.01) | — |
| B | ULV group | **-0.28 (-0.42 to -0.15)** | -0.07 (-0.20 to 0.06) |
|  | sULV group | **-0.21 (-0.36 to -0.07)** | — |
| C | ULV group | **-0.22 (-0.36 to -0.09)** | -0.06 (-0.19 to 0.07) |
|  | sULV group | -0.16 (-0.31 to -0.02) | — |
| Path efficiency (%) |  |  |  |
| A | ULV group | -6.36 (-16.86 to 4.14) | -7.08 (-17.26 to 3.10) |
|  | sULV group | 0.72 (-10.57 to 12.01) | — |
| B | ULV group | **-16.13 (-26.63 to -5.63)** | **-13.67 (-23.85 to -3.50)** |
|  | sULV group | -2.46 (-13.75 to 8.84) | — |
| C | ULV group | **-19.51 (-30.01 to -9.01)** | **-15.45 (-25.62 to -5.27)** |
|  | sULV group | -4.06 (-15.35 to 7.24) | — |
| Turn deviation |  |  |  |
| A | ULV group | 4.21 (-0.21 to 8.63) | 3.81 (-0.47 to 8.10) |
|  | sULV group | 0.40 (-4.35 to 5.15) | — |
| B | ULV group | **5.74 (1.32 to 10.16)** | 4.37 (0.09 to 8.65) |
|  | sULV group | 1.37 (-3.39 to 6.12) | — |
| C | ULV group | **7.80 (3.38 to 12.22)** | **6.06 (1.78 to 10.34)** |
|  | sULV group | 1.74 (-3.01 to 6.50) | — |

Significant mean differences (horizontal subtracted by vertical) are indicated in bold.

#### Metro Station

Group differences in motion onset latency, walking speed, path efficiency, and turn deviation across the three metro station environment wayfinding tasks are shown in Figure 7.

**Figure 7.**
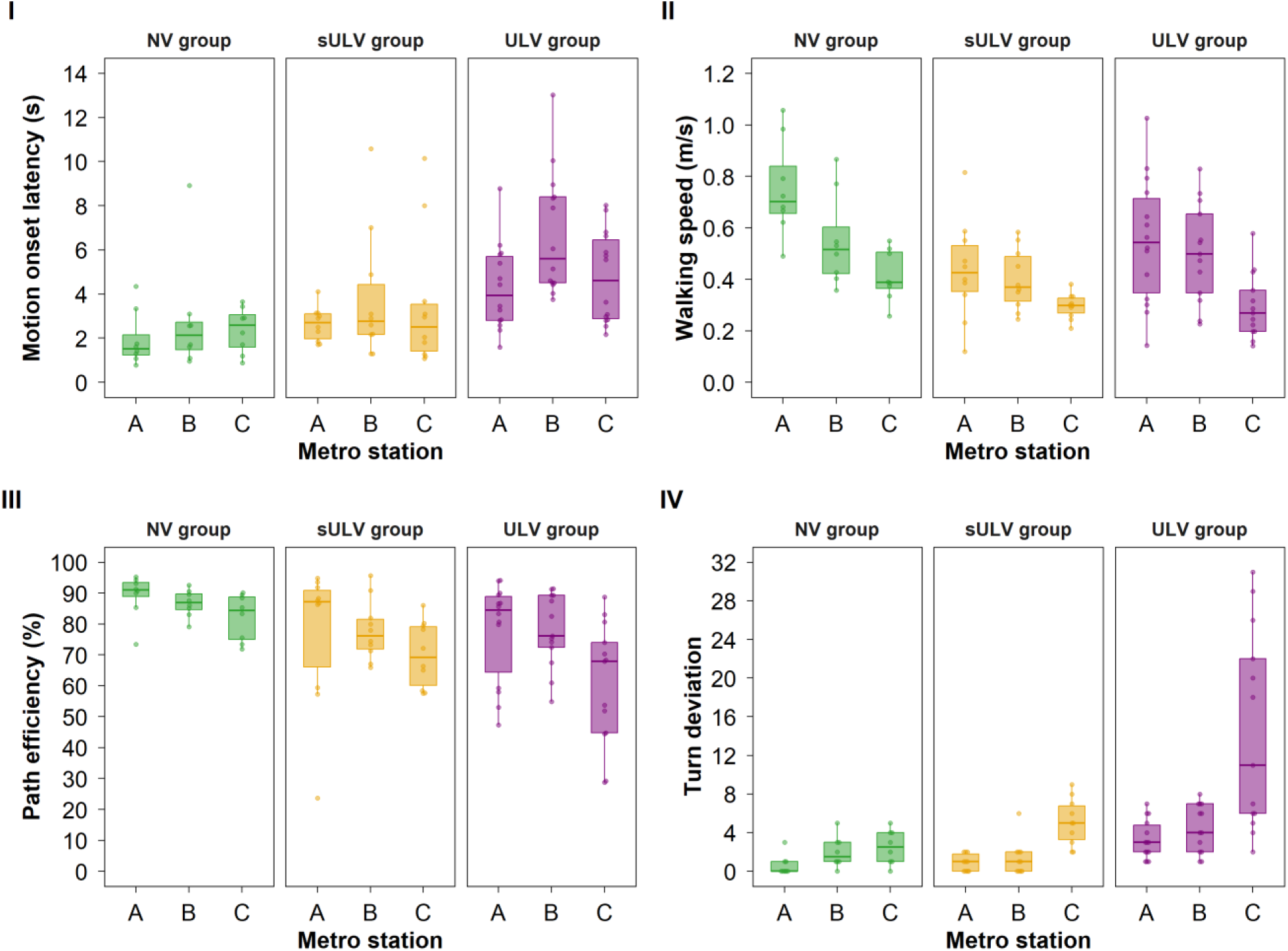
Box plots showing motion onset latency (I), walking speed (II), path efficiency (III) and turn deviations (IV) for the three metro station tasks (A. first level [walk to train entrance door], B. second level [walk to train entrance door and enter the train], and C. third level [walk to train entrance door, enter the train and identify an empty seat]) and the three groups (normal vision [NV], simulated ultra-low vision [sULV], and ultra-low vision [ULV]). Across all panels, box plots and overlaid data points use the same color, with NV shown in green, sULV in amber, and ULV in purple. The box indicates the interquartile range (25^th^-75^th^ percentile) and median; the whiskers indicate the minimum and maximum values excluding outliers.

In the metro station wayfinding tasks, a significant main effect of motion onset latency was found for participant group (F(2,87) = 8.19, p = 0.001), with the ULV group initiating movement 2.9 s and 1.9 s later than the NV group (5.2 vs. 2.4 s, approximately 2.2 times as long as the NV group, p < 0.001) and sULV group (p = 5.2 vs. 3.3 s, approximately 1.6 times as long, 0.007), respectively. There was also a significant main effect for level (F(2,87) = 5.07, p = 0.008), where level B (walk to train entrance door and enter the train) delayed walking initiation by 1.5 s compared with level A (walk to train entrance door) (4.4 vs. 3.0 s, approximately 1.5 times as long, p = 0.002). There were no significant interactions between NV, sULV and ULV group and level (F(4,87) = 0.92, p = 0.456, Figure 7.I).

For walking speed, there was a significant main effect of participant group (F(2,85) = 5.09, p < 0.001), with participants with sULV walking 0.20 m/s slower than those with NV (0.38 vs. 0.57 m/s, approximately 0.7 times the speed of the NV group, p = 0.002). There was also a significant level effect (F(2,85) = 25.56, p < 0.001), where the third level (level C) reduced walking speed by 0.25 m/s compared with the first level (level A) (0.33 vs. 0.58 m/s, approximately 0.6 times the speed, p < 0.001) and by 0.15 m/s compared with level B (0.33 vs. 0.49 m/s, approximately 0.7 times the speed, p < 0.001). No significant group by level interaction effect was observed (F(4,85) = 1.93, p = 0.113, Figure 7.II).

Path efficiency showed a significant main effect of participant group (F(2,85) = 5.03, p = 0.009), with the ULV group performing 13.8% less efficiently than the NV group (p = 0.003), respectively. A significant main effect of level was also observed (F(2,85) = 6.55, p = 0.002), where the path efficiency for level C was reduced by 10.3% compared with level A (p = 0.002) and by 10.0% compared with level B (p = 0.003). Again, no significant group by level interaction effect was found (F(4,85) = 1.01, p = 0.410, Figure 7.III).

Turn deviation showed significant main effects of participant group (F(2,85) = 12.65, p < 0.001) and level (F(2,85) = 19.08, p < 0.001), as well as a significant group by level interaction effect (F(4,85) = 5.75, p < 0.001, Figure 7.IV). Post-hoc pairwise analysis revealed that in the third level (level C), the ULV group made 12 more turns than the NV group (p < 0.001) and 9 more than the sULV group (p < 0.001), while no significant difference in turn deviation was found between the NV and the sULV groups (p = 0.203). No significant differences were found between levels A and B across all three participant groups (p > 0.005).

Generalized linear mixed models results for all outcome measures of metro station environment wayfinding tasks, including F values, degrees of freedom, and p value, are provided in Table 6. Bonferroni-adjusted post-hoc pairwise comparisons between the three participant groups for all metro station environment outcome measures are summarized in Table 7.

**Table 6.** Generalized linear mixed models for all the outcome measures in the metro station environment wayfinding tasks.

| Outcome measures | Effects | F value | df1 | df2 | p value |
| --- | --- | --- | --- | --- | --- |
| Motion onset latency | Group | 8.19 | 2 | 87 | **0.001** |
|  | Level | 5.07 | 2 | 87 | **0.008** |
|  | Group X Level | 0.92 | 4 | 87 | 0.456 |
| Walking speed | Group | 5.09 | 2 | 85 | **< 0.001** |
|  | Level | 25.56 | 2 | 85 | **< 0.001** |
|  | Group X Level | 1.93 | 4 | 85 | 0.113 |
| Path efficiency | Group | 5.03 | 2 | 85 | **0.009** |
|  | Level | 6.55 | 2 | 85 | **0.002** |
|  | Group X Level | 1.01 | 4 | 85 | 0.410 |
| Turn deviation | Group | 12.65 | 2 | 85 | **< 0.001** |
|  | Level | 19.08 | 2 | 85 | **< 0.001** |
|  | Group X Level | 5.75 | 4 | 85 | **< 0.001** |

Significant p values are indicated in bold.

**Table 7.** Bonferroni-adjusted post-hoc pairwise comparisons between the three groups for all outcome measures in the metro station environment.

|  |  | Mean difference (95% confidence interval) |  |
| --- | --- | --- | --- |
| Motion onset latency (s) |  | NV group | sULV group |
| A | ULV group | 2.34 (0.35 to 4.33) | 1.65 (-0.20 to 3.51) |
|  | sULV group | 0.69 (-1.44 to 2.81) | — |
| B | ULV group | **3.88 (1.89 to 5.86)** | **2.89 (1.04 to 4.75)** |
|  | sULV group | 0.98 (-1.14 to 3.11) | — |
| C | ULV group | 2.38 (0.40 to 4.37) | 1.23 (-0.63 to 3.08) |
|  | sULV group | 1.16 (-0.97 to 3.28) | — |
| Walking speed (m/s) |  |  |  |
| A | ULV group | **-0.20 (-0.35 to -0.05)** | 0.11 (-0.03 to 0.25) |
|  | sULV group | **-0.32 (-0.48 to -0.16)** | — |
| B | ULV group | -0.04 (-0.19 to 0.11) | 0.11 (-0.03 to 0.25) |
|  | sULV group | -0.15 (-0.31 to 0.01) | — |
| C | ULV group | -0.12 (-0.28 to 0.03) | -0.01 (-0.15 to 0.14) |
|  | sULV group | -0.12 (-0.28 to 0.04) | — |
| Path efficiency (%) |  |  |  |
| A | ULV group | -11.49 (-24.07 to 1.09) | 0.77 (-10.99 to 12.52) |
|  | sULV group | -12.26 (-25.72 to 1.21) | — |
| B | ULV group | -8.23 (-20.97 to 4.51) | 0.64 (-11.29 to 12.56) |
|  | sULV group | -8.87 (-22.33 to 4.60) | — |
| C | ULV group | **-21.59 (-34.33 to -8.85)** | -9.50 (-21.42 to 2.43) |
|  | sULV group | -12.09 (-25.56 to 1.37) | — |
| Turn deviation |  |  |  |
| A | ULV group | 2.66 (-1.11 to 6.43) | 2.39 (-1.13 to 5.90) |
|  | sULV group | 0.28 (-3.76 to 4.31) | — |
| B | ULV group | 2.29 (-1.52 to 6.11) | 2.89 (-0.68 to 6.47) |
|  | sULV group | -0.60 (-4.63 to 3.43) | — |
| C | ULV group | **11.94 (8.12 to 15.75)** | **9.34 (5.77 to 12.91)** |
|  | sULV group | 2.60 (-1.43 to 6.63) | — |

Significant mean differences (horizontal subtracted by vertical) are indicated in bold.

### Comparison across virtual reality environments – street crossing vs. cafeteria vs. metro station wayfinding tasks

Outcome measure data were collapsed across complexity levels, excluding level A of the street crossing environment (no obstacles; straight-ahead walking), and wayfinding performance was compared across the street crossing, cafeteria, and metro station environments within each participant group. The comparisons of wayfinding performance across VR environments for each participant group is shown in Table 8.

**Table 8.** Comparisons of wayfinding performance across virtual reality environments for each participant group.

| **Ultra-low vision group** | **F value** | **df1** | **df2** | **p value** |
| --- | --- | --- | --- | --- |
| Motion onset latency | 3.46 | 2 | 26 | **0.047** |
| Walking speed* | 20.98 | 1.4 | 17.8 | **< 0.001** |
| Path efficiency | 10.43 | 2 | 26 | **< 0.001** |
| Turn deviation | 1.52 | 2 | 26 | 0.237 |
| **Simulated ultra-low vision group** |  |  |  |  |
| Motion onset latency | 7.10 | 2 | 18 | **0.005** |
| Walking speed | 11.36 | 2 | 18 | **0.001** |
| Path efficiency | 30.39 | 2 | 18 | **< 0.001** |
| Turn deviation | 0.56 | 2 | 18 | 0.582 |
| **Normal vision group** |  |  |  |  |
| Motion onset latency | 2.90 | 2 | 14 | 0.088 |
| Walking speed | 7.12 | 2 | 14 | **0.007** |
| Path efficiency | 8.47 | 2 | 14 | **0.004** |
| Turn deviation | 0.47 | 2 | 14 | 0.635 |

F values and p values from repeated measures analysis of variance. Sphericity was assessed using Mauchly's test, with violations ( $p > 0.05$ ) indicated by \*.
Significant p values are indicated in bold.

In the ULV group, environment had a significant effect on motion onset latency (F(2,26) = 3.46, p = 0.046), walking speed (F(1.4,17.8) = 20.98, p < 0.001), and path efficiency (F(2,26) = 10.43, p < 0.001). Post-hoc pairwise analysis showed metro station environment reduced walking speed by 0.17 m/s (0.45 vs. 0.63 m/s, approximately 0.7 times the speed, p = 0.003) and path efficiency by 11.4% (72.9 vs. 84.3%; p = 0.003) compared to the street crossing environment. The cafeteria environment also reduced walking speed by 0.21 m/s (0.42 vs. 0.63 m/s, approximately 0.7 times the speed, p < 0.001) compared to the street crossing environment.

Similarly, in the sULV group, environment had a significant effect on motion onset latency (F(2,18) = 7.10, p = 0.005), walking speed (F(2,18) = 11.36, p = 0.001) and path efficiency (F(2,18) = 30.39, p < 0.001). Post-hoc analysis showed that the street crossing environment delayed walking initiation by 3.1 s compared with the cafeteria environment (6.2 vs. 3.1 s, approximately 2.0 times as long, p = 0.049) and by 2.9 s compared with the metro station environment (6.2 vs. 3.3 s, approximately 1.9 times as long, p = 0.036). The metro station environment reduced walking speed by 0.20 m/s (0.38 vs. 0.57 m/s, approximately 0.7 times the speed, p = 0.002) compared to street crossing environment. Again, the metro station environment reduced path efficiency by 15.0% (75.0 vs. 90.0%, p = 0.001) compared with the street crossing environment, and by 15.3% (75.0 vs. 90.3%, p = 0.001) compared with the cafeteria environment.

In the NV group, environment had a significant effect on walking speed (F(2,14) = 7.12, p = 0.007) and path efficiency (F(2,14) = 8.47, p = 0.004). Post-hoc analysis showed that the metro station environment reduced path efficiency by 7.0% (86.1 vs. 93.1%, p = 0.049) compared with the street crossing environment, and by 7.0% (86.1 vs. 93.1%, p = 0.016) compared with the cafeteria environment.

### Correlation between trajectory-based metrics and visual acuity in participants with ultra-low vision

Visual inspection of the data indicated greater overall variability in navigation performance among participants with ULV compared to the sULV and NV groups. To explore this variance, correlation analyses were conducted within the ULV group between visual acuity and trajectory-based metrics. Visual acuity showed a significant positive correlation only with motion onset latency in the metro station environment (r = 0.58, p = 0.038) indicating that worse visual acuity was associated with longer initiation. Notably, visual acuity did not significantly correlate with walking speed, path efficiency, or turn deviation across any of the three environments.

## DISCUSSION

This study used a calibrated VR wayfinding platform to characterize navigation behavior in individuals with ULV across street crossing, cafeteria, and metro station environments of increasing complexity, and to compare their performance with the NV and sULV groups. The findings demonstrate that visual wayfinding difficulty in ULV varied across environments rather than remaining uniform, and that trajectory-based metrics were effective at capturing these differences. Participants with ULV generally showed longer motion onset latency, lower walking speed, reduced path efficiency, and greater turn deviation than participants with NV, with participants with sULV exhibiting performance levels between those of the ULV and NV groups. Among the tested environments, the metro station posed the greatest wayfinding challenge. Path efficiency appeared to be the most informative outcome measure, as it consistently captured differences across environments and reflected overall navigation behavior, while turn deviation provided complementary information in more complex tasks. Taken together, these findings highlight that visual wayfinding involves multiple interacting components and cannot be fully described using a single outcome measure.

A notable observation was that different environments appeared to challenge distinct aspects of navigation behavior during wayfinding. In the street crossing environment, increasing complexity with moving cars (levels C and D) was associated with worse wayfinding performance across all measures, particularly in participants with ULV. These changes suggest that dynamic traffic environments increase uncertainty and promote more cautious behavior. This interpretation is consistent with prior evidence showing that navigation under degraded viewing conditions imposes added attentional demands, as individuals must monitor safety while moving and process environmental information at the same time (Rand, Creem-Regehr, & Thompson, 2015). Accordingly, motion onset latency in the present study may not reflect motor initiation as much as hesitation and decision-making difficulty before starting to move under visually uncertain conditions. From this perspective, navigation difficulty in the street crossing environment may have reflected challenges in action timing, hazard anticipation, and confidence in traffic gap judgments, rather than route planning alone.

In contrast, the cafeteria environment appeared to challenge route organization and spatial updating more strongly. Increasing clutter delayed motion onset in ULV and substantially reduced path efficiency relative to both NV and sULV. These findings suggest that visually cluttered indoor environments may disrupt efficient route planning and updating, even when the destination is relatively near. More generally, wayfinding depends not only on obstacle detection but also on spatial updating, route memory, and the integration of self-motion cues over time, and these processes can lead to systematic errors as environments become more complex (Loomis et al., 1993). Prior work on spatial updating during navigation further shows that people must continuously update their position and direction as they navigate, particularly when visual information is degraded (Rieser, Guth, & Hill, 1986). The cafeteria task likely challenged this updating process by requiring frequent reorientation around irregularly arranged obstacles and visually salient distractors.

The metro station environment imposed the greatest wayfinding difficulty, as shown at the most complex level (level C). At this level, turn deviation increased substantially in ULV. Across-environment analyses further showed that the metro station consistently reduced path efficiency in the ULV, sULV, and NV groups. Unlike the street crossing or cafeteria, the metro environment required sequential goal completion across multiple stages, with each stage involving reorientation and route correction in the presence of dynamic social elements. Prior work has shown that navigation difficulty can increase disproportionately when tasks require multiple spatial transformations and updates in sequence, even in relatively small-scale environments (Loomis et al., 1993; Rieser, Guth, & Hill, 1986). The combination of multi-step planning, repeated reorientation, and crowd interaction in the metro station environment likely amplified small inefficiencies in perception and spatial updating, explaining why it emerged as the most challenging setting overall.

An important implication of these findings is that path efficiency and RDP-derived turn deviation may be particularly informative for assessing visual wayfinding difficulty. Notably, we observed greater variability in navigation performance among participants with ULV. Standard clinical vision measures do not fully explain this variance; as our results show, while visual acuity correlated with onset latency in the metro station environment, it did not predict active routing behaviors like path efficiency or turn deviation. Consequently, participants with similar residual vision exhibited different navigation behaviors. This is clinically relevant because conventional mobility outcomes, such as completion time, obstacle collision, walking speed, or percentage of preferred walking speed, along with clinical vision measures, may not fully capture how an individual navigates through an environment (Chang, Dillon, Deverell, Boon, & Keay, 2020; Virgili & Rubin, 2010). An individual may complete a task at a similar overall speed yet take a more indirect route, hesitate repeatedly, or require multiple route corrections. In the present study, path efficiency and turn deviation appeared to capture changes in route and directional control more effectively than speed measures alone, supporting the value of trajectory-based metrics for assessing functional navigation. This is particularly valuable for future rehabilitation strategies; for example, while visual implants restore limited vision and cannot be easily altered, VR-based tools can quantify navigation behavior to maximize functional vision by identifying specific, trainable navigation skills.

Walking speed differed across participant groups and levels, but in the present study it was calculated differently from conventional mobility-course measures. Instead of using a predefined course length, walking speed was based on the actual distance walked and the active task duration (after removing motion onset latency) derived from continuous head position data. Therefore, walking speed reflected movement speed after walking initiation rather than overall task speed. Because walking speed was based on the actual distance walked rather than a fixed course length, it was more sensitive to hesitation, detours, and less direct navigation in complex environments. This further highlights the value of VR-based trajectory analysis for assessing navigation behavior.

Another key observation was that sULV only partially reproduced the navigation behavior observed in ULV. Participants with sULV often walked more slowly than those with NV and, in some settings, also showed reduced path efficiency. Within the sULV group, the street crossing environment produced the longest motion onset latency, suggesting that sudden simulated vision loss may particularly delay movement initiation in safety-critical, dynamic environments where participants must judge when it is safe to begin walking. In contrast, the metro station environment reduced walking speed and path efficiency, suggesting greater difficulty during active navigation once movement had begun. However, compared with NV, sULV did not consistently reproduce the larger inefficiencies and excess turning observed in ULV across environments. This suggests that short-term blur simulation captures some consequences of reduced visual input but not the full behavioral profile associated with native ULV. These differences may reflect additional factors, such as long-term adaptation to profound vision loss, altered visual search and scanning strategies, greater reliance on nonvisual cues, and increased uncertainty in interpreting residual vision. Accordingly, while sULV remains a useful comparison condition, it should not be assumed to fully model the navigational experience of individuals with ULV.

The present study has several methodological strengths, including the comparison of three ecologically relevant environments within a common VR framework, the use of graded levels of complexity, and the inclusion of ULV, sULV, and NV groups. Navigation behavior was quantified using continuous head-tracking data and trajectory-derived outcomes, extending beyond the feasibility and validity of many earlier VR mobility studies (Aleman et al., 2021; Authie et al., 2024; Bennett et al., 2023). The VR wayfinding instrument was also designed to include factors such as high contrast, high luminance, and brighter lighting, which are important for visual wayfinding in individuals with ULV who rely on rudimentary vision (Adeyemo et al., 2017; Kartha, Sadeghi, Swanson, & Dagnelie, 2022). Good reproducibility of optimal path marking further supports the path efficiency and turn deviation analysis. Together, these features demonstrate the value of VR as a safe and controlled way to study navigation behavior and compare how different environments and outcome measures reveal wayfinding difficulty.

The study has several limitations. The environments and complexity levels were presented sequentially, so order or fatigue effects cannot be ruled out. The sULV condition was informative, but it cannot fully represent those who live with ULV. In addition, navigation measures were derived from head position rather than full-body movement. Although VR offers strong control and safety, it cannot fully reproduce the sensory (auditory and haptic cues), and contextual demands of real-world travel. The study also did not report head rotation pattern, which could be particularly informative for understanding how individuals with ULV make traffic gap judgments in the street crossing environment.

In conclusion, these findings show that visual wayfinding difficulty in ULV is strongly influenced by environmental context. Within this VR paradigm, the metro station posed the greatest challenge overall, and path efficiency as well as turn deviation emerged as the most sensitive behavioral measures of environmental difficulty. These results support the use of VR-based trajectory analysis for studying functional navigation in people with profound vision loss and suggest that future mobility assessment in ULV may benefit from emphasizing route-based measures alongside conventional ones. More broadly, this approach may help identify the environments and behavioral features that are most informative for evaluating functional vision and guiding future rehabilitation strategies in people with profound vision loss.

## Data Availability

All data produced in the present study are available upon reasonable request to the authors

